# Multiple Instance Learning Framework can Facilitate Explainability in Murmur Detection

**DOI:** 10.1101/2022.12.08.22283240

**Authors:** Maurice Rohr, Benedikt Müller, Sebastian Dill, Gökhan Güney, Christoph Hoog Antink

**Affiliations:** KIS*MED – AI Systems in Medicine, Technische Universität Darmstadt, Darmstadt, Germany

## Abstract

**Objective:** Cardiovascular diseases (CVDs) account for a high fatality rate worldwide. Heart murmurs can be detected from phonocardiograms (PCGs) and may indicate CVDs. Still they are often overlooked as their detection and correct clinical interpretation requires expert skills. In this work, we aim to predict the presence of murmurs and clinical outcome from multiple PCG recordings employing an explainable multitask model.

**Approach:** Our approach consists of a two-stage multitask model. In the first stage, we predict the murmur presence in single PCGs using a multiple instance learning (MIL) framework. MIL also allows us to derive sample-wise classifications (i.e. murmur locations) while only needing one annotation per recording (“weak label”) during training. In the second stage, we fuse explainable hand-crafted features with features from a pooling-based artificial neural network (PANN) derived from the MIL framework. Finally, we predict the presence of murmurs as well as the clinical outcome for a single patient based on multiple recordings using a simple feed-forward neural network.

**Main results:** We show qualitatively and quantitatively that the MIL approach yields useful features and can be used to detect murmurs on multiple time instances and may thus guide a practitioner through PCGs. We analyze the second stage of the model in terms of murmur classification and clinical outcome. We achieved a weighted accuracy of 0.714 and an outcome cost of 13612 when using the PANN model and demographic features on the CirCor dataset (hidden testset of the George B. Moody PhysioNet challenge 2022, team “Heart2Beat”, rank 12 / 40).

**Significance:** To the best of our knowledge, we are the first to demonstrate the usefulness of MIL in PCG classification. Also, we showcase how the explainability of the model can be analyzed quantitatively, thus avoiding confirmation bias inherent to many post-hoc methods. Finally, our overall results demonstrate the merit of employing MIL combined with handcrafted features for the generation of explainable features as well as for a competitive classification performance.

## 1. Introduction

Cardiovascular diseases (CVDs) are considered to make up more than a third of all deaths globally per year [1]. Therefore, screening for CVDs is of public interest. While there exists a multitude of mandatory preventive medical examinations and techniques, such as heart auscultation, blood pressure measurement, imaging methods, electrocardiography, and even genetic testing, the availability and ease of use is not always given. Phonocardiograms (PCGs), which are recordings of all sounds of the heart during a cardiac cycle, are widely available and allow physicians to replay measurements and, after digital processing, to visually inspect these recordings. Also, they enable further algorithmic processing and potentially automated diagnosis.

Heart murmurs are audible vibrations caused by perturbations of the blood flow such as strong pressure gradients or velocity changes and are often indicators for CVDs. For this reason, the most recent George B. Moody PhysioNet challenge 2022 [2, 3] raised the task of detecting heart murmurs in PCG recordings from four different auscultation locations of a pediatric population and predicting the clinical outcome of a full diagnostic screening. For children, this is particularly interesting because 80 % of children have heart murmurs at the early stage of their life [4]. As the presence and predictive capability for CVDs of the murmur varies with age, periodic screening is recommended. But when it comes to the recognition of heart murmurs via auscultation, highly skilled doctors with good hearing capabilities [5] are required. Due to the general shortage of doctors, this extends to need for AI support.

Polls show, year after year, that while the trust in AI systems in the general population increases, the overall trust is comparably low for diagnostic medical systems [6]. However, in order to bring machine learning models to practice, physicians and patients have to be convinced of their trustworthiness. This is evident for example in recent policy making in Germany, where the importance of explainability in decision support systems for medicine is emphasized and even made a requirement that has to be documented [7].

At the same time, frequently used explainablity methods come with several drawbacks. For example, so-called posthoc methods like saliency maps, SHAP and LIME, tend to be susceptible to confirmation bias, because users (and even researchers) can interpret the output of the explainer favorably towards their model [8]. This is aggravated by the fact that while the model performance is quantitatively analyzed via a wide range of metrics, analysis of the explainability is usually only done qualitatively and not actually checked for performance. Recent publications show first results of the application of explainable methods to PCG classification employing SHAP [9] or Attention mechanisms [10]. A different kind of explainability methods is called *Prototypical explanations* [8, 11], which can be considered as a form of inherent explainability. Here, the model is trained not only to classify samples but to also identify prototypical elements of each class. This makes it possible to quantify how much of each component is identified for a given decision.

Besides their structure, approaches to murmur detection differ in three important processing steps: Preprocessing, use of segmentation, integration of feature extraction and / or deep learning. For preprocessing we found large differences in recent publications. Some authors used heavy preprocessing by down-sampling to 1 kHz and limiting the frequency band to 25–400 Hz or less [12–16] or down-sampling to 2 kHz instead [17], others employed no filtering and immediately transformed the recordings into time-frequency domain [18, 19], while showing similar prediction accuracy. Heart cycle based segmentation of the PCG was found to be important if hand-crafted features are used [20, 21]. For deep learning based models and large datasets segmentation seems to be not as relevant [17, 22]. Especially in the George B. Moody PhysioNet challenge 2022, hierarchical or multi-stage models were popular due to the variability in possible features in the data and the task of predicting both the presence of murmur and the clinical outcome for the patient.

In this work, we propose a 2-stage multitask model that predicts the presence of murmur in a PCG recording (stage 1) and then combines recordings from multiple locations in an inference step with demographic information in order to predict the clinical outcome as well as murmur probability for the patient (stage 2). We reason that in order to detect murmur from a single recording, the recording by itself is sufficient, and information such as age is only of interest in the second stage where we apply a shallow model. We focus on explainability of the model output between those two stages by integrating a model inspired by the multiple instance learning (MIL) framework [23] and handcrafted features, as this is where insights can be useful to help medical professionals which is generally neglected.

Therefore, we first discuss the dataset (2.1), the general preprocessing and the data-augmentation (2.2). We then motivate our multiple-instance learning approach and show how we arrive at the notion that it provides good features (2.3). We supplement these features with more handcrafted features which are also meaningful (2.4). Finally we put the parts together to the full 2-stage model (2.5.2).

## 2. Methods

We propose a new model architecture for murmur enhancement and detection that benefits from self-learned features, but also includes segmentation-based features associated with certain murmur characteristics (Section 2.4). The general approach is shown in Figure 1. After preprocessing the PCGs for training, realistic data-augmentation is performed. We then interpret the detection of heart murmurs as a two stage multiple instance learning problem (further explained in Section 2.3) with strong labels at the sample level and at the recording level which can be exploited at the respective stages. The Pooling-based artificial neural network (PANN) is derived from the MIL inspired U-Net [24] (MIL-U-Net) model that shows that instance-wise features can be reasonably assumed. For evaluation, the PANN models prediction performance on single recordings is shown. The PANN features are then combined in the second stage with demographic and the segmentation-based features in a simple fully connected neural network that predicts the presence of murmur in a patient as well as the medical outcome for the same patient. The methods are evaluated on public datasets listed in Section 2.1.

**Figure 1.**
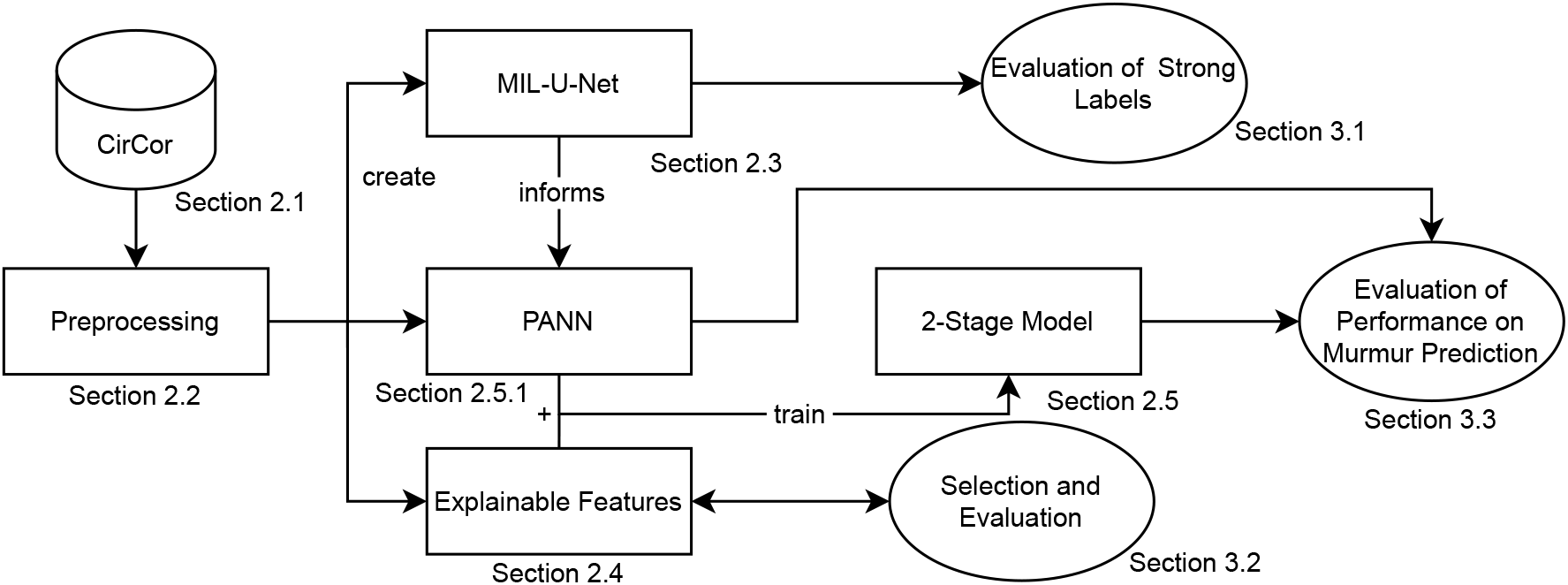
The general approach as described in this paper

### 2.1. Datasets

We use two public datasets to qualitatively and quantitatively analyze our proposed methodology.

#### CirCor Dataset

The Círculo do Coração (Heart Circle - CirCor) Dataset was used in the George Moodey PhysioNet Challenge 2022 and contains a total number of 5282 heart sound recordings from the main four auscultation locations of 1568 patients aged between 0.1 and 356.1 months. Most participants are in the 1-11 year age group. For a more detailed description of the dataset, we refer to the original description in [25, 26]. A segmentation into the phases of the heart cycle (S1, systole, S2, diastole) was created algorithmically and verified by experts. Murmurs are classified in terms of their timing (early-, mid-, and late-systolic/diastolic) (Table 2), shape, pitch, quality, and grade. Each patient is also labeled as “murmur present / unknown / absent” by an expert annotator and “clinical outcome normal / abnormal” from the assessment of a pediatric cardiologist. The label distribution is shown in Table 1. It is important to note that the clinical outcome label is not directly connected to the murmur label.

**Table 1.** CirCor training set label distribution of clinical outcome and presence of murmur in patients

| **Murmur / Outcome** | Normal | Abnormal | $\Sigma$ |
| --- | --- | --- | --- |
| Present | 29 | 150 | 179 |
| Unknown | 25 | 43 | 68 |
| Absent | 432 | 263 | 695 |
| $\Sigma$ | 486 | 456 | 942 |

**Table 2.** CirCor training set murmur timings in PCG recordings

| Phase / Timing | Early | Mid | Late | Holo | Total |
| --- | --- | --- | --- | --- | --- |
| Systolic | 143 | 56 | 2 | 291 | 492 |
| Diastolic | 14 | 0 | 0 | 0 | 14 |
| Systolic & Diastolic | - | - | - | - | 12 |

The data is divided into training (60 %), validation and test set. For model cross-validation we only had direct access to the training data.

#### MHSDB

[27] The “Michigan Heart Sound and Murmur Database” consist of 24 recordings with different types of heart sounds and murmurs. The database is aimed at training doctors in recognizing different heart sounds. Thus the recordings are very clean and the heart sounds and murmurs are easily identifiable.

### 2.2. Preprocessing

Firstly, we resample the recordings to 2000 Hz using linear interpolation. We compute the signal quality by applying the two criteria as described by Plesinger et al. [28] which are similar to those used by Springer et al. [29] to identify low noise signals: In short, a segment of a signal is considered saturated if high and low values are over-represented in its amplitude histogram. A segment is considered too noisy if the energy of the Hilbert envelope of the 100 to 250 Hz bandpassed signal is higher than that of the 15 to 90 Hz bandpassed signal. The signal is subsequently filtered with a 10^th^ order Butterworth band-pass filter with cut-off frequency of 10 to 800 Hz. For training of the models we choose the segment of the signal with the highest signal quality. Signals with less than three recorded usable heart cycles are discarded.

#### 2.2.1. Augmentation

Due to the relatively small size of the training dataset, we employ data augmentation. During training and after preprocessing, one or multiple augmentations are performed at random: *scaling, Gaussian noise, drop, cutout, shift, resampling, random resampling, sine wave addition, bandpass filtering*. Scaling randomly rescales the signal. Gaussian noise adds Gaussian noise to the signal. Drop randomly sets signal values to zero. Cutout randomly sets signal intervals to zero. Shift randomly shifts the signal in time (creating zeros at either end). Random resampling uses random time-warping to simulate a changing heart rate. Resampling linearly resamples the signal to another sampling frequency, simulating another heart rate. Sine wave adds a random sine wave to the signal. Bandpass filtering randomly applies a bandpass filter between 0.2 and 45 Hz. The probabilities for each augmentation were defined based on the empirical impact of each individual augmentation on the models validation performance as presented in [30].

### 2.3. MIL-U-Net

The detection of heart murmurs can be formulated as machine learning problem with so-called “weakly labeled” data, as known from sound event detection [31]. These types of problems can be solved using the *multiple instance learning* (MIL) framework while not relying on prior segmentation. Weakly labeled in this context means that for each sound recording, only a single tag (e.g. “murmur present”) is provided without knowing the exact onset and offset times of the relevant section in the recording. More specifically, in MIL [23, 31], a weakly labeled dataset *D* = *{B*_*n*_, **y**_*n*_*}*^*N*^ consists of a set of bags 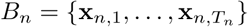, where each bag is a collection of instances. *T*_*n*_ is the number of instances in the *n*^th^ bag and each instance has a length (duration) *d*. Given a particular sound class *k*, a bag (PCG recording) is considered *positive* (e.g. “murmur present”, *y*_*k,n*_ = 1) if it contains at least one positive instance and *negative* (*y*_*k,n*_ = 0) if it contains no positive instance at all. The problem formulation heavily implies that, given a prediction for each instance of a bag, the condensed label is given by the maximum of these predictions. McFee et al. [31] argue that while in some cases the maximum operator is indeed the best option to condense the output of multiple instances, it also comes with two drawbacks: First, the output of a model applying this operator is only dependent on a single output and thus learning is highly unstable and sensitive to initialization; Second, it leads to favoring features of the signal that have the best predictive power, while ignoring features that are also predictive but not as strongly. While averaging the predictions for each instance instead would circumvent all of these problems, it does not take the main assumption behind MIL into account and thus gives too much responsibility to instances with little information. One solution to these problems is combining the feature of giving maximum responsibility to the most important instance while still leaving some weight on the other instances. Softmax-Pooling

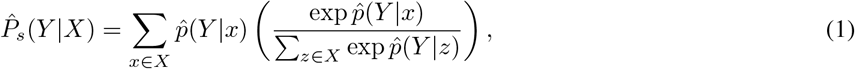

where 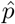 is the likelihood of an instance being classified as *y* and 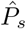 is the likelihood of the bag being classified as *y* as the result, weights each instance prediction between [0, 1] such that the most prominent instance has the largest impact.

In order to predict the instance probability 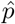 (also termed “murmurness”), we employ a simple U-Net architecture [24] with rectified linear unit (ReLU) activations, 5 downsampling steps and a kernel size of 5 as depicted in Figure 2. Since the U-Net is a fully convolutional network, we can circumvent having to decide the size of an instance and efficiently compute an instance level prediction for each sample. The size of an instance is learned by the network.

**Figure 2.**
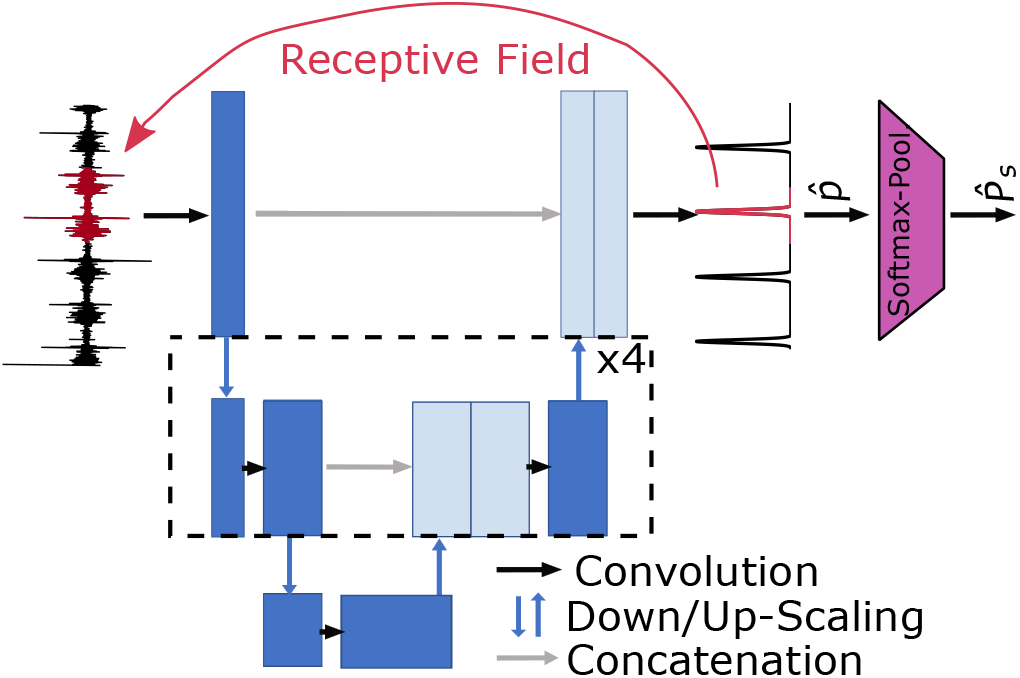
MIL-U-Net Model for murmur detection

#### 2.3.1. Training

We train the network on the complete official CirCor training set employing 10-fold-cross-validation on the patient level. We hold out 20 % of the 9 training folds for validation during training and use early stopping. As cost function, we use binary cross entropy and optimize with Adam, with a batch size of 128, a learning rate of 0.001 and decreasing it every 10 epochs by a factor of 0.5. For more efficient training, we also fix the signal-length to 2^14^ samples and zero-pad the signal if necessary. The output signals of the validation folds are then stored for further processing.

#### 2.3.2. Postprocessing

The activation of the network varies strongly in relevant areas. Therefore, after applying a sigmoid function, we threshold the signal at 0.6 and find the strongest peaks using a peak finding algorithm. If the peaks are within a margin of 60 ms they are considered as stemming from the same murmur instance. The peaks are merged, resulting in a segmentation as predicted in Figure 3.

**Figure 3.**
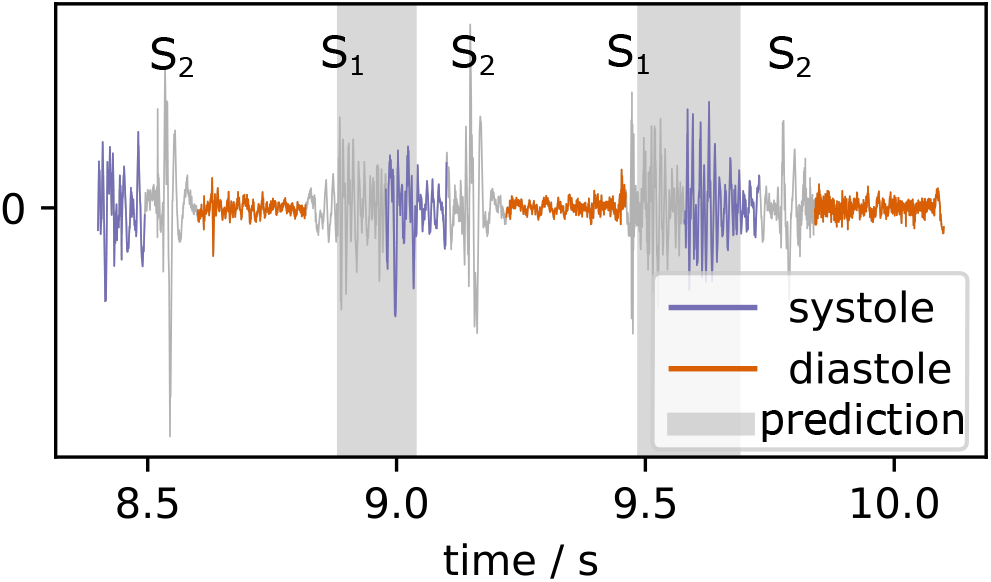
Example of postprocessed MIL-U-Net output for a patient diagnosed with systolic murmur (CirCor recording 9979 AV)

#### 2.3.3. Quantitative Explainability Analysis

In order to evaluate the detection performance of MIL-U-Net at instance level, we first apply it to a selection of murmur and non-murmur recordings from MHSDB. In a second evaluation, we use the CirCor dataset and segment all PCG recordings into the heart phases. To verify the results, we also use reference segmentations from the dataset itself. We assume that if a recording is labeled as “murmur not hearable”, all instances are without murmur and label them as 0. If a recording is labeled as “murmur hearable”, we look at the timing assigned by the doctor, which could mean systolic, diastolic or both. For simplicity we also assume, that murmur occurs in every heart cycle for the whole length of each phase. If the segmentation by the network overlaps with the respective murmur phase, we count it as true positive otherwise as false positive.

### 2.4. Segmentation-based Explainable Features

The generation of segmentation-based explainable features follows the general idea of first creating a variety of features that is then reduced using feature importance and correlation analysis. The murmur classification task is taken as the basis to calculate the feature importance and results in 22 features starting from a set of 622 features. Our model is then extended by these features that can be interpreted by themselves to some extent.

Due to the higher sensitivity to signal distortions of the explainable features, additional preprocessing is performed before feature extraction. Noisy fragments that often appear at the begin and the end of recordings, presumably due to positioning of the stethoscope, are removed. Thus, we linearly increase the cut off part from 0 s at 5 s recording length to 3 s at 30 seconds recording length. For despiking we use the algorithm proposed by Schmidt et al. [32]. Afterwards, we only consider the longest parts of the signal, for whose Hilbert envelope *Ŝ*_e_ holds

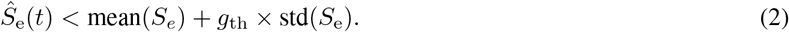

In this work, *g*_th_ was set to 2.7 empirically.

The start and the end of the resulting signal are also clipped if they are longer than 5 seconds, as they are close to the thresholds. This ensures that strong disturbances are removed and only sequences of high quality are kept. We calculate segmentations for S1, S2, diastole and systole using the segmentation algorithm of Springer et al. [33].

The features can be formally categorized as demographic / anthropometric features, duration features, time-frequency features and shape features.

The **5** demographic / anthropometric features are age, sex, weight, height, weight/height.

The duration features are computed as the averages of the durations of S1, S2, diastole, systole, full heart cycle, and the ratios of all these durations which sums up to **15** features.

The time-frequency domain features (Figure 4) rely on the amplitudes of the preprocessed signal, 4 transformations of the signal which are the spectral centroid, spectral bandwidth [34], signal energy computed in a window of 13 ms and Hilbert envelopes computed for the frequency bands LF (15 to 90 Hz), MF (55 to 150 Hz), HF (100 to 250) Hz, SF (200 to 450 Hz) and UF (400 to 800 Hz) as proposed by Plesinger et al. [28]. Additional signals were generated by dividing the bandlimited Hilbert envelopes by each other, e.g. the envelopes for HF/LF, HF/MF, MF/LF, SF/MF, SF/LF, UF/LF, and (HF+MF)/LF, resulting in 7 new signal variants. For each of these signal transformations we computed the mean, standard deviation, and maximum value over the whole signal and for each of the four heart phases and full cycle. We compute the ratio of these values. This results in (3 + 15 + 15) *×* 18 = 594 features.

**Figure 4.**
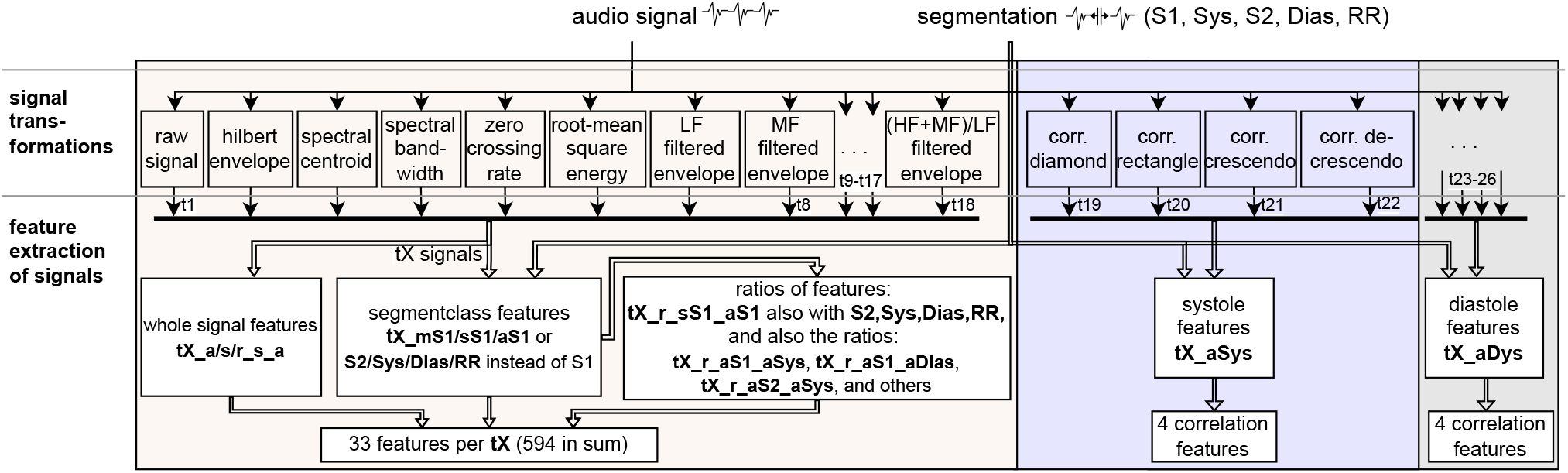
Generation process of time-frequency domain and shape features. The feature names are abbreviated as: a = average, s = standard derviation, m = maximum, r = ratio of two signals, tX = X^th^ signal transformation as outlined above. These result in 602 features, which together with the demographic- (5) and segment duration based features (15) sum up to 622.

Additionally we compute **8** shape features based on the most prominent murmur shapes which are described as diamond shaped, plateau shaped, crescendo and decrescendo. Based on the average duration of systole and diastole we create adapted templates for each that are stretched over the whole duration.

This process results in 5 + 15 + 594 + 8 = **622** features in total. However, the features are not equally descriptive of heart murmur as we expect features that compare systole to heart cycle to be most important. High frequency content compared to the normal heart sounds could also be indicative of murmur. In order to reduce the number of features, we train an XGBoost model [35] on a simple binary classification problem, where we only consider recordings with the same labels as described in Section 2.3. We then utilize the inbuilt feature importance metric to rank all 622 features. From these 622 features, we select the 20 most important for further analysis.

### 2.5. 2-stage Pooling-based NN

We adapt the main MIL-U-Net idea of pooling neural network features and create the *pooling-based artificial neural network* (PANN) (Figure 5). The PANN is used in the first stage for low dimensional feature generation and location selection that are fused together with demographic features and features from Section 2.4 in the second stage multi-label model. The models predicts the clinical outcome and the presence of murmur for a single patient based on multiple recordings.

**Figure 5.**
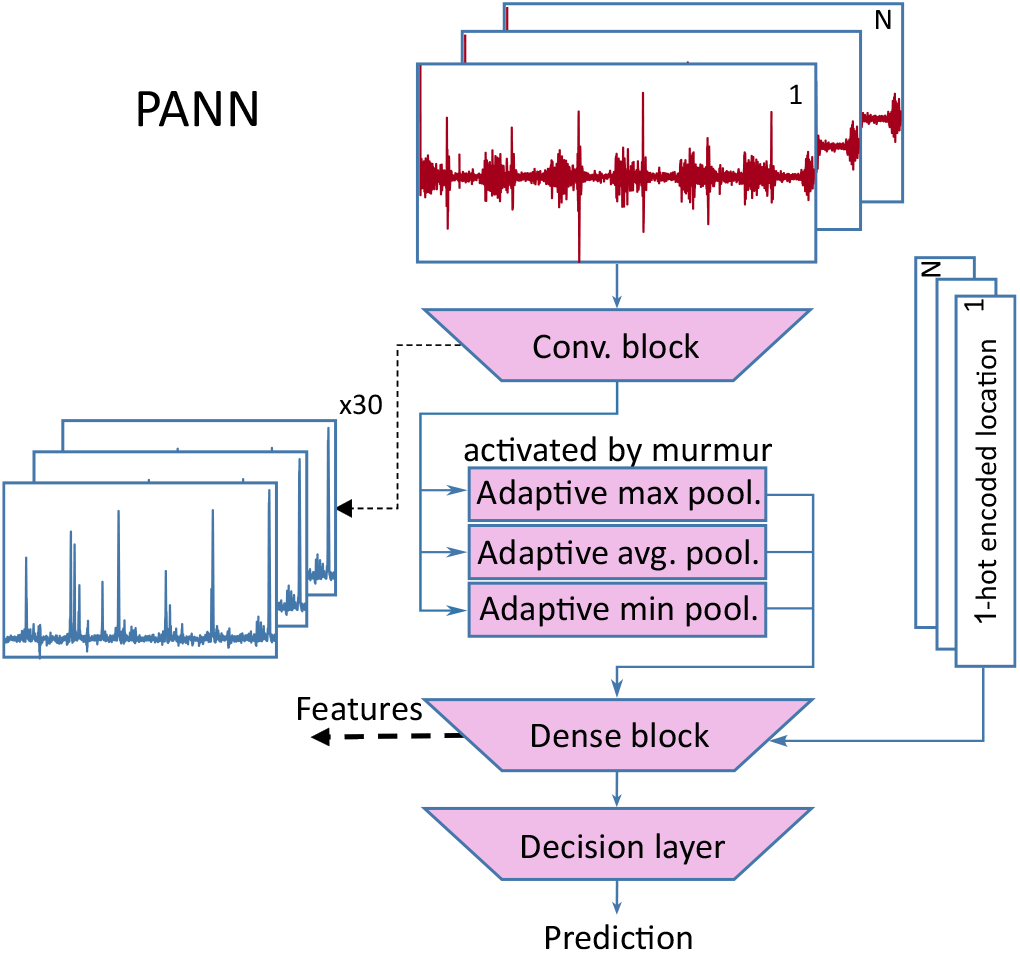
Pooling based artificial neural network for feature generation

#### 2.5.1. Pooling-based Artificial Neural Network (1st stage)

The PANN employs a convolutional encoder consisting of 6 base blocks. The base block firsts computes 1d-convolutions (kernel size=5, padding=same), then batch norm, another 1d-convolution of the same type, dropout (rate=0.1) and max pooling (stride=2) to encode the signals in a feature-rich representation. These features resemble the signal output of the MIL-U-Net, but are less explainable because they are processed by different adaptive pooling layers that produce a fixed-sized output of 30 features in total. 15 of those features are computed by an adaptive max-pooling layer, 10 features are computed by an adaptive average-pooling layer, and 5 are computed by an adaptive min-pooling layer. The combination of max- and average-pooling has a similar effect as the softmax-pooling layer in Figure 2. By dividing the signal into multiple parts that are pooled, we can reduce the effect of noisy segments of the signal on the pooling output since later stages of the processing can still learn to compensate for inconclusive outputs. These output features of the pooling layer are then combined in a fully connected layer (30 *×* 64 + 5 input, 100 hidden and 20 output neurons) with the one-hot encoded recording locations. These outputs are then evaluated in a linear decision layer with softmax activation function. By processing each recording location separately and combining the detection results based on simple decision rules, we enable the user to verify the suspected murmur position.

#### 2.5.2. Multi-label model (2nd stage)

The second stage is a simple feed forward neural network that is fed with precomputed features which include (1) the 20 most important features as described in Section 2.4, (2) the encoded output of the PANN model and its location wise predictions, as well as (3) demographic features such as age-group and weight. The model consists of 5 feed forward layers with (123,492,246,20) hidden neurons, each followed by batch norm and leaky ReLU activations. Dropout is only used after these hidden layers. The output layer is made up of 3 neurons for murmur prediction and 2 for outcome prediction. In both cases, a softmax activation computes the conditional probability.

#### 2.5.3. Training

The PANN model is trained first on single recordings of CirCor splitting the data in a stratified way based on the murmur label of each patient. Each recording is then labeled as “Unknown” if the patient’s murmur label is “Unknown”, “Present” if murmur is hearable in the recording, and “Absent” otherwise. The training optimizes the cross entropy loss. The weights (*w*_m_, *w*_o_) for the cost function are computed as the inverse relative occurrence of each class to counteract class imbalance.

The learning rate is set to 0.001 initially and scheduling reduces the learning rate by a factor of 0.3 after each 30 epochs. The best model is picked based on the mean of training and validation accuracy. Data augmentation is employed extensively using the presented methods to facilitate generalization beyond the training data. After this procedure, the trained model is only used as a feature generator. For the second stage, all features (1, 2, 3) are precomputed for each patient. As before, for each output weighted cross entropy loss is employed,

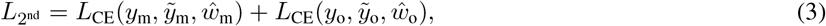

where *L*_CE_ is the cross-entropy loss, *y*_m_, *y*_o_ are the murmur / outcome labels and 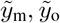 the murmur / outcome prediction respectively. The weights are adjusted based on the factors in the weighted accuracy metric [2], where *ŵ*_o_ = *w*_o_[1, 5]^*T*^ (Normal, Abnormal) and *ŵ*_m_ = *w*_m_[1, 3, 5]^*T*^ (Absent, Unknown, Present).

### 2.6. Metrics

We use AUROC, AUPRC, F1, accuracy and the metrics defined by [2], which are weighted accuracy and outcome cost, to validate our model. The weighted accuracy is defined as

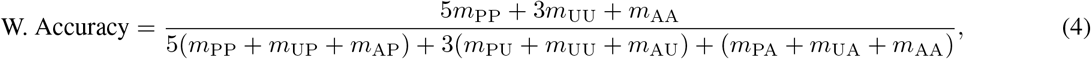

where *m*_PP_, *m*_UU_, *m*_AA_ are the number of correctly predicted labels “Present”, “Unknown”, “Absent” respectively. *m*_*P U*_ is the number of examples predicted as “Unknown” but labeled “Present” and so forth [2].

The outcome cost is defined as

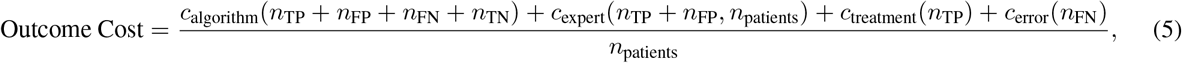

where *c*_*i*_ are four cost functions incorporating the algorithmic cost, the cost of a medical expert, the cost for the treatment and the cost of an untreated disease. The functions are each dependent on the number of true positive *n*_TP_, false positive *n*_FP_, false negative *n*_FN_, true negative *n*_TN_ predictions, and the total number of patients *n*_patients_. Additional information can be found in [2].

## 3. Results

We evaluate the MIL approach first as it motivates the PANN model. We then show the feature selection process leading to 22 features that are plugged into the 2-Stage model of Section 2.5.2. Finally, the performance of the 2-Stage model is compared with recently published models.

### 3.1. MIL Approach

We analyzed the performance of the MIL-U-Net in a 5-fold-cross-validation on *CirCor*. The prediction accuracy for murmur on a per-sound-recording basis achieved an average of 0.784. This result is not discussed in more detail, as it qualitatively shows that the network learns plausible features for murmur detection and the focus of the MIL approach lies on the generation of good “strong labels” for the subsequent model.

Next, we inspected the predictions for recordings of the *MHSDB* and the respective activation qualitatively as depicted in Figure 6. If there is no murmur in the recording the resulting prediction is “0” and thus the activation of the network (murmurness) for all specified samples is also close to 0. The model segments systolic murmur reliably in multiple examples. Normal heart sounds do not lead to spikes in the murmurness signal in general and in the normal PCG no false positives are detected. The murmurness is highest during systole, while abnormal PCG features like missing S2 sounds can lead to wider regions of murmurness spikes. Diastolic murmur does not lead to strong activations in the inspected examples.

**Figure 6.**
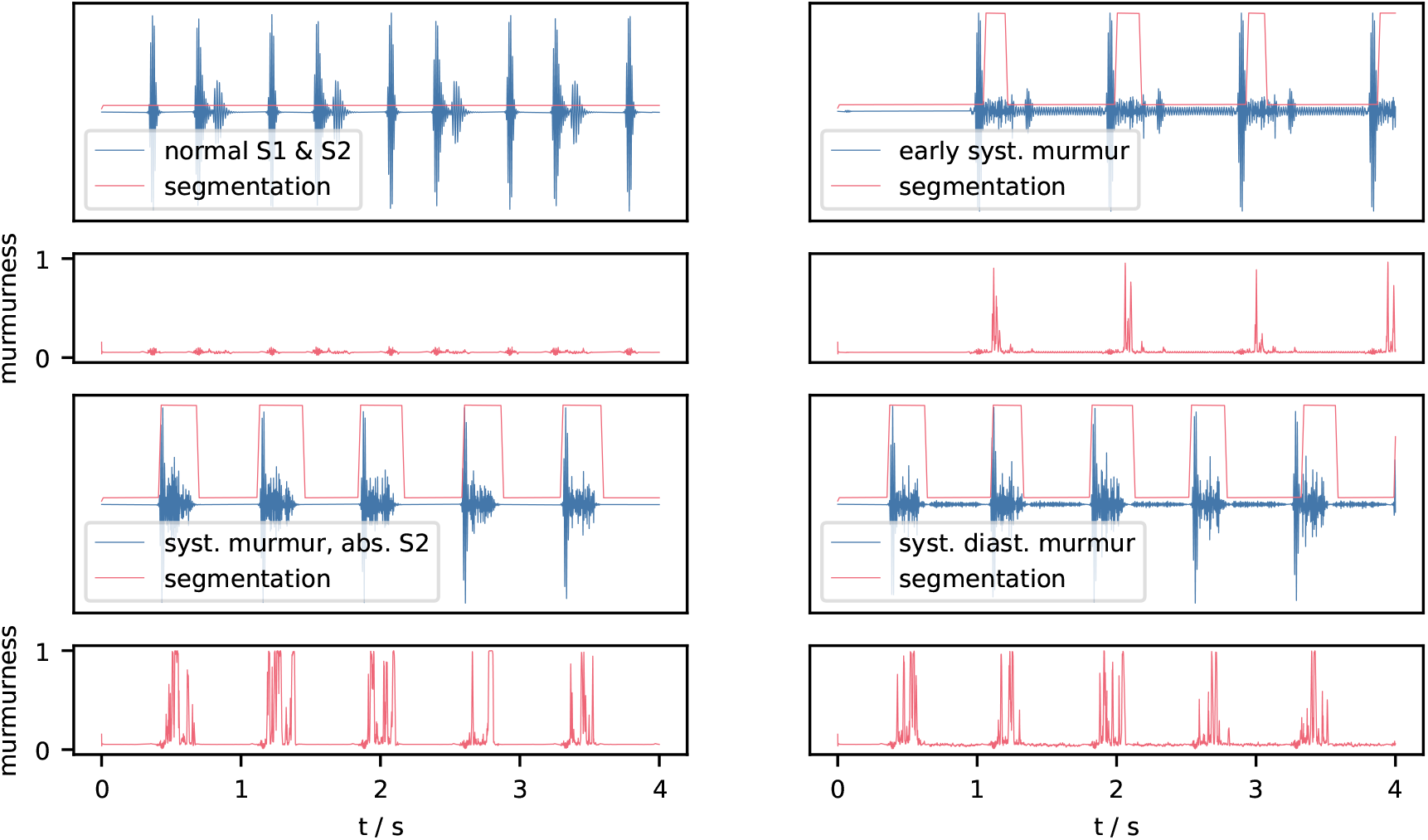
Postprocessed segmentation of MIL-U-Net (red) overlayed on the respective audio signal (blue), the activation of the the network termed “murmurness” is shown below each recording.

Quantitative results were generated by joining the results of the cross-validation and analyzing the post-processed activations of the MIL-U-Net in the respective heart-cycle phases as depicted in Figure 7. The simple assumption is that if a signal contains murmur, it probably is present in the same phase per heart cycle. We therefore computed the precision for each recording for each phase by dividing the number of activations that lie in the respective phase by the total number of activations. If no activation was counted, the whole signal was marked as “no activation”. For comparison and to detect potential bias, we also provide the pseudo precision achieved if systolic murmur was assumed for recordings without murmur. Here no activation is expected. For recordings whose murmur timing was labeled as “systole”, the precision for murmur detection shows a strong tendency to be close to 1.0. This mostly includes signals with 20-50 distinct activations. We can cross-check this result by looking at pseudo precision achieved in the systole for non-murmur recordings. It does not show the same distribution and more importantly high precision values are not standing out. Mostly no activation is registered. For murmur timing labeled as diastole few examples are available, but a similar trend to systolic murmur can be observed. Recordings without murmur appear to generate mostly false activations, but these are mostly singular events.

**Figure 7.**
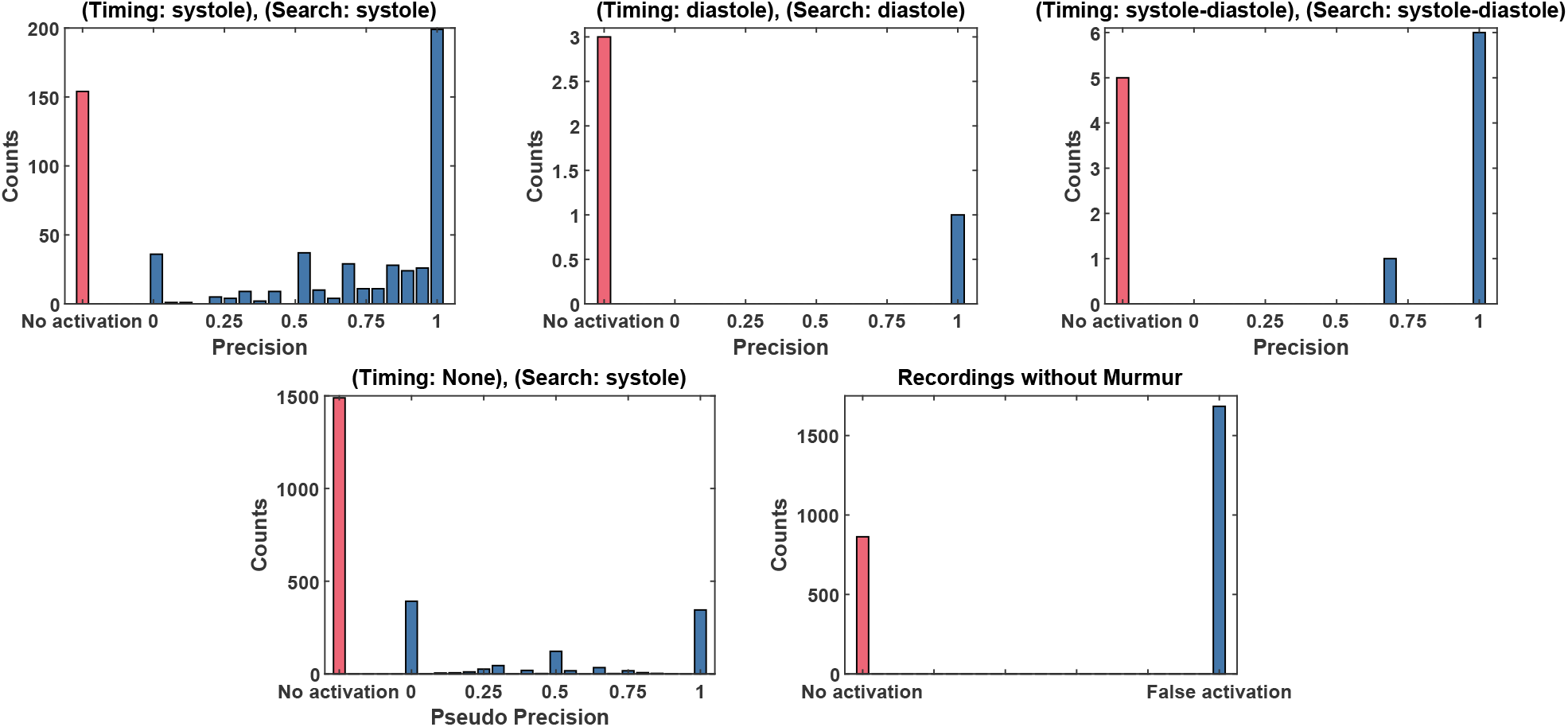
Virtual murmur segmentation performance on CirCor of MIL-U-Net when considering activation in diastole/systole as hit for diastolic/systolic murmur. Recordings without activation are marked as red. For comparison the bottom left plot shows shows the pseudo precision if we assume recordings without murmur to contain systolic murmur.

### 3.2. Feature Selection

We evaluated the XGBoost model in a 5-fold cross-validation using the weighted accuracy score on murmur classification for the full set of features as well as for the best 20 features, the best 10 and best 3 features. Since the model using only the best 20 features was within an error margin of 1 % to the model using the full set of features and 10 features resulted in 4 % reduced score, we continued with the best 20 features. A simple statistical correlation analysis between the feature values and the respective labels yields a similar result, but the inter-correlation of the features impedes the selection based on correlation alone.

The demographic features showed no relevance for the murmur detection task, but since the model predicts murmur and clinical outcome simultaneously, we consider the features *weight* and *age*. Both showed a comparably high importance for the clinical outcome decision. Therefore, this step results in the selection of 22 features listed in Table 3 to be used in the final model. Most features put a high relevance on high frequency signal contents and the relationship between the signal in the systole compared to the rest of the heart cycle. The top 20 features showed in parts also direct correlation with the murmur label. The demographic features showed no relevance for the murmur detection task. We included them nonetheless because our final model predicts murmur and clinical outcome simultaneously and age and weight showed a comparably high importance for the clinical outcome decision.

**Table 3.**
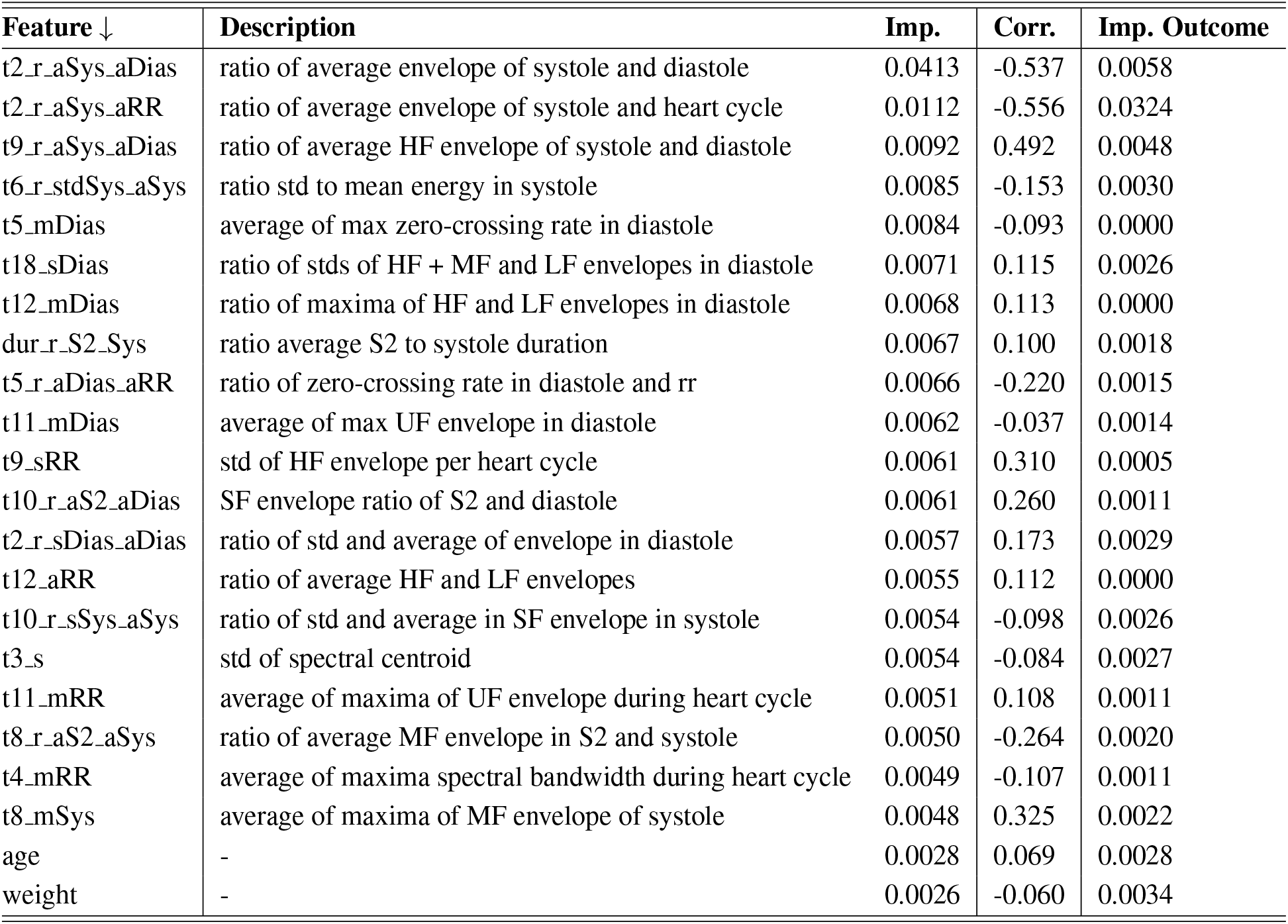
Top 20 selected features plus weight and age and their respective feature importance values (regarding murmur presence and Clinical Outcome) and correlation with murmur presence

### 3.3. Model Performance

The final model performance for the two-stage model was evaluated using a 10-fold cross-validation and f1-score, accuracy, AUROC, AUPRC and weighted accuracy for murmur classification and the outcome cost function as well as accuracy, f1-score, AUROC, AUPRC for the medical outcome classification. The results in Table 4 imply that adding the handcrafted features to the deep learning model does not improve performance. Table 5 shows the results for the murmur classification task of the model without handcrafted features in context of the challenge on the hidden test. Table 6 shows the respective outcome score for our team.

**Table 4.** Results of 10-fold cross-validation for murmur detection using both PANN features and explainable features and only PANN features as average (standard deviation).

| model (murmur) | AUROC | AUPRC | F1 | Accuracy | W. Accuracy |
| --- | --- | --- | --- | --- | --- |
| PANN + explainable features | 0.804 (0.047) | 0.604 (0.025) | 0.532 (0.034) | 0.726 (0.051) | 0.704 (0.050) |
| PANN features + weight & age | 0.833 (0.068) | 0.664 (0.094) | 0.533 (0.105) | 0.718 (0.084) | 0.714 (0.099) |
| model (outcome) | AUROC | AUPRC | F1 | Accuracy | Outcome Cost |
| PANN + explainable features | 0.646 (0.048) | 0.0654 (0.036) | 0.5794 (0.065) | 0.586 (0.064) | 13948 (1011) |
| PANN features + weight & age | 0.652 (0.035) | 0.0656 (0.019) | 0.596 (0.052) | 0.601 (0.053) | 13612 (1975) |

**Table 5.**
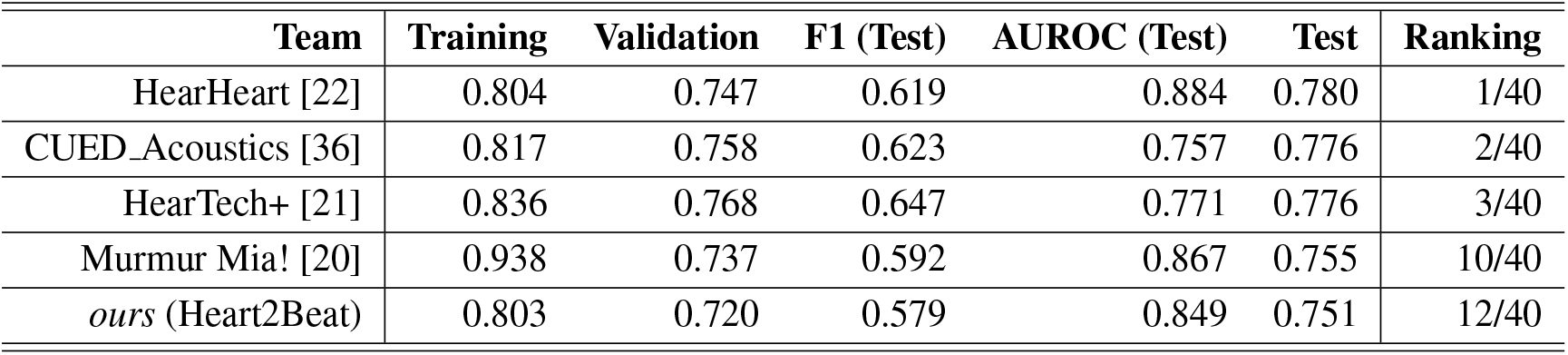
Weighted accuracy metric scores (official challenge score) for our final selected entry (team Heart2Beat) for the murmur detection task, including the ranking of our team on the hidden test set. For the test set we also provide F1 and AUROC.

**Table 6.** Cost metric scores (official challenge score) for our final selected entry (team Heart2Beat) for the clinical outcome identification task, including the ranking of our team on the hidden test set.

| Training | Validation | Test | Ranking |
| --- | --- | --- | --- |
| 10810 | 9135 | 12244 | 11/39 |

## 4. Discussion

The multiple instance learning framework can be shown to work in principle for murmur detection and leads to plausible murmur segmentations. The qualitative analysis of the generated segmentations shows promising results for our approach, even though it only relies on the assumption of the existence of strong labels for the whole PCG. Because the maximum operator is a crucial element, only a single example of murmur in a recording suffices to create a good weak label. The data shows that even though globally the models prediction is correct, certain instances occur to be labeled wrongly. This seems to be immanent to the approach. The precision statistics for *murmurness* validate that a meaningful murmur detection can be learned via MIL and the indicated positions are often accurate. Robustness is not yet achieved and is our goal in future work. Signals without murmur usually show only little false activations if any. The results can thus be seen as a prove of concept that needs further refinement. Especially diastolic murmur does not lead to strong activation which is most probably due to the under-representation in the training dataset and more data would definitely help. By translating the idea of stronger labels on instance basis to the classification task, good results can be achieved as shown in Table 5. The Softmax-Pooling layer is a promising step that should also be implemented in an adaptive fashion.

The feature selection process confirmed that demographic features are not relevant for the pure prediction of heart murmur. We also compared the distributions of all features for both recordings containing murmur and not containing murmur in the feature selection process. Better performing features usually show less overlap in their distributions and thus, the correlation with the output label is higher. We tried the best 5, 10, 15, 20, 25, 30, and all features during model training. The best 20 performed the best with regards to the weighted accuracy metric. Features showing higher frequency content in systole compared to the rest of the signal are assigned high feature importance, which goes in line with the notion that this is a strong indicator for heart murmur. Features comparing the envelope of systole and diastole are also assigned high feature importance which might be due to the bias towards systolic murmur in the CirCor dataset. Also including weight and age for the outcome prediction made a difference. Nonetheless, only including weight and age with the PANN features produced a better performing model than including the handcrafted explainable features. We also found that not training both optimization goals simultaneously improves the results for murmur prediction, which could be inferred from the fact that in the dataset both outcome and murmur presence were not closely related. What also comes as a surprise is that using the whole signals instead of signals where strong noise is removed during inference did not seem to make a difference for prediction. That could be interesting further research. The PANN learning stability is sub-optimal so far and can only be approached by doing a cross-validation for model selection. Better methods such as replacing the max-pooling must be evaluated. The final 2-stage model achieved competitive results in the George B. Moody Challenge. Comparing the results of the different approaches on training, validation and test sets, we see a high variability for all models, which shows that relying on the metrics from a single dataset might be misleading.

## 5. Conclusion

We evaluated the MIL framework as an approach to create explainable features for murmur detection. We quantified the explanatory power by computing the the precision of MIL-U-Net’s instance-level predictions and thus presented a method to quantify explainability, which is dearly needed. The result shows that the model’s predictions are mostly reasonable, as long as there is sufficient training data. Selectively looking at results were the model performed well or badly, and checking the features shows that post-hoc explanations, where we as researches tend to blame false predictions on noisy parts or positive predictions on certain aspects of the signal, are always near-by. Therefore it is beneficial, that the MIL-U-Net can provide a segmentation of the signal that can be used to simplify double-checking the prediction by an expert.

Still, explanations drawn from the MIL approach and the handcrafted features can introduce harmful biases into decision making [8], because for some PCG recordings it was more difficult to make sense of the model output. Analyzing alleged explanations statistically, as we did, therefore seems to be a more suitable approach.

The overall model performance is in the range of the top performing models for both outcome and murmur classification. Thus showing that utilizing MIL combined with handcrafted features is a suitable approach not only for generating explanations but also achieving competitive classification performance.

## Data Availability

All data produced in the present study are available upon reasonable request to the authors.

https://physionet.org/content/circor-heart-sound/

https://open.umich.edu/find/open-educational-resources/medical/heart-sound-murmur-library

## Notes

### Competing Interest Statement

The authors have declared no competing interest.

### Funding Statement

This study did not receive any funding

### Author Declarations

https://physionet.org/content/circor-heart-sound/ and https://open.umich.edu/find/open-educational-resources/medical/heart-sound-murmur-library

